# Plasmin Cleavage of Beta-2-Glycoprotein I Alters its Structure and Ability to Bind to Pathogenic Antibodies

**DOI:** 10.1101/2024.04.12.24305747

**Authors:** Hannah F. Bradford, Christophe J. Lalaurie, Jayesh Gor, Xin Gao, Charis Pericleous, Stephen J. Perkins, Hannah Britt, Konstantinos Thalassinos, Ian Giles, Anisur Rahman, Mihaela Delcea, Paul A. Dalby, Thomas C.R. McDonnell

## Abstract

Beta-2-Glycoprotein I (β2GPI) is the main autoantigenic target of antiphospholipid syndrome (APS) with antibodies leading to clinical manifestations. There are two known structural isomers of β2GPI, a J shape and a circular shaped one. The transition between these structures is incompletely understood, with the functional implications unknown. β2GPI is a substrate of the protease plasmin, which cleaves within the fifth domain of β2GPI leading to altered cellular binding. Very little is currently known regarding the structure and function of this protein variant. We present the first comprehensive structural characterisation plasmin-clipped β2GPI and the associated implications for pathogenic antibody binding to this protein.

**Methods:** β2GPI was purified using a novel acid-free process from healthy control plasma and cleaved with plasmin. Cleavage was confirmed by SDS-PAGE. Structural characterisation was undertaken using dynamic light scattering (DLS), small angle X-ray scattering (SAXS), ion mobility mass spectrometry (IMMS) and molecular dynamics simulation (MD). Activity was tested using inhibition of β2GPI ELISAs with patient samples and cleaved β2GPI in the fluid phase and cellular binding by flow cytometry using HUVEC cells.

**Results:** DLS revealed a significantly smaller hydrodynamic radius for plasmin-clipped β2GPI (p=0.0043). SAXS and MD analysis indicated a novel S-like structure of β2GPI only present in the plasmin-clipped sample whilst IMMS showed a different structure distributions in plasmin clipped compared to non-clipped B2GPI. The increased binding of autoantibodies was shown for plasmin-clipped β2GPI (p=0.056), implying a greater exposure of pathogenic epitopes following cleavage.

**Conclusions:** Cleavage of β2GPI by plasmin results in the production of a unique S-shaped structural conformation and higher patient antibody binding. This novel structure may explain the loss of binding to phospholipids and increase in anti-angiogenic potential described previously for plasmin-clipped β2GPI.

## Introduction

Beta-2-Glycoprotein I (β2GPI) is a serum glycoprotein of approximately 50 kDa, comprised of four structurally similar domains (DI-DIV) and a structurally distinct fifth domain (DV) which are arranged in a single chain like beads on a string. β2GPI circulates at a concentration of approximately 200 μg/ml and has to date been proposed to exist in two possible structures an “open” J-shaped and “circular” O-shaped structure (1). The first four domains are typical members of the short complement regulator (SCR) superfamily, termed domains DI-DIV while the C terminal domain (DV) contains lysine-rich regions on the surface, responsible for cellular binding. The activities of β2GPI are widely varying from angiogenesis to complement activation, whilst the structure of β2GPI is hypothesised to have significant implications for the activity of the molecule based on the assumption that, in the circular form, various motifs are hidden from solvent, thus preventing interactions with expected binding partners (1).

In circulation, β2GPI has a number of contrasting functions and is almost unique (2) in being capable of both up- and down-regulation of activation of the complement and coagulation cascades. It is incompletely understood how these opposing functions are balanced; however, it has been hypothesised that regulation between these opposing functions is through structural restriction. Therefore, the structure of β2GPI, and the factors that modify it, are crucial to fully understand its activity and function.

The structure of β2GPI is complex, and as previously mentioned it can form a linear (J-shaped; PDB ID: 1C1Z) or a circular (O-shaped) structure. However, it has been suggested that β2GPI can also form an intermediate or alternative structure which is S-shaped, as first shown by Hammel et al using small angle X-ray scattering (SAXS) (3–7). The regulation of its structure is incompletely understood, however, several factors have been shown *in vitro* to trigger structural change, including changes in pH, salt concentration (1), disulphide reduction (8) and lysine acetylation (9). Genetic manipulations to delete a disulphide to mimic the impact of reduction (10), although less clinically relevant, has also been shown by SAXS to alter β2GPI structure reverting to the J shape rather than the O-shape. Most recently Kumar et al. studied the potential for a shift in serum concentration to purely linear β2GPI, highlighting the potential for this equilibrium to be shifted and altering the exposure of protein motifs (11).

The role of β2GPI in complement and coagulation is further complicated by the fact that it is a substrate for plasmin. β2GPI plays a role in the conversion of plasminogen to plasmin, facilitating this through the binding of the fifth domain to plasminogen (12). The cleavage of β2GPI by plasmin takes place between the eighth and ninth amino acids from the C-terminus, yielding a peptide by-product of eight amino acids, and a shortened fifth domain. This modification of β2GPI prevents its binding to plasminogen and thus slows the conversion of plasminogen to plasmin in the absence of other cofactors for plasminogen cleavage (13). This loss of activity suggests a change in binding and function after cleavage, which has been shown in other studies (14–16).

Plasmin-clipped β2GPI can act as an inhibitor of angiogenesis (14), a function much less prominent in the non-cleaved form. Furthermore, the cleavage of β2GPI leads to increased cardiac manifestations in neonatal lupus *in vivo* (17) due to the loss of the binding site for Ro60, an autoantigen linked to other autoimmune disorders which functions as an RNA scavenger. β2GPI binds in the central pore of Ro60, preventing the binding of anti-Ro60 autoantibodies. The Anti-Ro60/Ro60 complex induces foetal heart block, therefore β2GPI binding rescues this phenotype, plasmin cleavage disrupts the 5^th^ domain (DV) of β2GPI and thus prevents binding to Ro60. Plasmin-clipped β2GPI has also been linked to cerebral infarction in antiphospholipid syndrome (APS) patients (16), whilst un-cleaved β2GPI has not been shown to have these same effects. Notably, Itoh et al. (18) showed increased levels of plasmin-clipped B2GPI in leukaemia patients, specifically associated with increased thrombosis. Despite this physiological role, the structures adopted by plasmin-clipped β2GPI are yet to be elucidated.

β2GPI is also the main auto-antigenic target of Antiphospholipid Syndrome (APS). APS is an autoimmune clotting disorder which is the leading cause of strokes under 50 years old and recurrent miscarriage. Patients routinely develop autoantibodies to Cardiolipin and β2GPI, both of which feature in diagnostic clinical criteria, however, a number of non-criteria antibodies also exist. Several studies have placed significant importance on anti-B2GPI autoantibodies, both diagnostically and mechanistically, as these antibodies are pathogenic in a range of mouse models (19,20). There is a polyclonal response with anti-B2GPI autoantibodies formed against a wide range of epitopes, however the two dominant sites for autoreactivity are DI and DV. Within the 1^st^ domain, a cryptic neo-epitope has been defined (R39-G43) to which the most pathogenic anti-β2GPI antibodies are formed, which have been shown to be thrombotic both *in vitro* and *in vivo*. Reversal agents which specifically target DI (21,22) have reduced clotting in *in vitro* functional assays and in both acute and chronic APS mouse models (23–25). The formation of the anti-β2GPI/β2GPI complex can trigger thrombosis in several ways, and notably in mouse models of APS, complement is also required to develop thrombosis, as C3−/− phenotypes rescue the mouse from thrombosis (26). The influence of β2GPI structure in APS is less well known, with few structural methods applied to β2GPI from patient serum. The circular form of B2GPI, traditionally hypothesised to be dominant in serum, theoretically hides both the DI and DV epitopes, implying the requirement of an open β2GPI form for antibody complex formation and thus disease progression (27). This has been refuted by recent research (11). The role of plasmin cleavage in APS is even less well studied and given the role of β2GPI in regulating both complement and coagulation, and the role of these cascades in APS, a naturally occurring cleavage by those cascades on a regulator is interesting. For example, during thrombosis β2GPI may act as a co-factor increasing this process, in doing so also by proxy activating complement and cleaving β2GPI which then loses the ability to regulate other functions. As such it is vital to understand the structures formed by β2GPI post plasmin cleavage to study the effect of plasmin clipped β2GPI in APS.

Here, we use a combination of biophysical and *in silico* techniques to characterise the structures of β2GPI generated by plasmin cleavage, and in order to gain a better understanding of how the structures are formed and stabilised in solution. The identification of novel structures may lead to an increased understanding of autoantibody generation, thrombosis formation and how the structure of β2GPI affects its role in various bodily systems.

## Methods

### Purification of β2GPI

Plasma was separated by SepMate (StemCell, Cambridge, UK). 50 ml of venous blood (collected in 10 ml sodium heparin vacutainers) was diluted 1:1 with RPMI 1640 media, meanwhile, 15 ml of Ficoll-Paque was dispensed into a 50 ml SepMate tube and 33 ml of diluted blood gently layered on top. Samples were centrifuged at 1,200 × g for 10 minutes and the top layer was poured into a 50 ml tube and re-spun for 5 minutes at 500 × g. Supernatant was again poured off and stored at −20 °C before use. β2GPI was precipitated using a polyethylene glycol (PEG) precipitation method as previously described (28). Briefly, plasma was diluted 1:4 with 10 mM sodium phosphate (pH 6.8) and 40% PEG 4000 (VWR, Lutterworth, UK) in 10 mM sodium phosphate (pH 6.8) was cooled on ice. The 40% PEG 4000 solution was then added drop wise to the diluted plasma, whilst mixing on a magnetic plate. For 35 ml of plasma, 12.5 ml of 10 mM sodium phosphate was added and a further 75 ml of PEG 4000 to a final concentration of PEG 4000 of 25%. This mix was then incubated at 4 °C for 30 minutes. Precipitate was collected in 50 ml Falcon tubes by centrifugation at 3000 × g for 30 minutes. Supernatant was discarded and precipitate was re-solubilised in 20 mM Tris pH 8.0, 30 mM NaCl at an appropriate volume. This was centrifuged for 30 minutes at 3000 × g to remove insoluble matter and the supernatant taken forward to purification. Purification was initially across 3 × 1 ml Heparin FF columns (Cytiva, Buckingham, UK) at a flow rate of 1 ml/min. Samples were pump loaded and the pressure was kept below 0.3 mPa. Once loaded the column was washed with 30 ml of 20 mM Tris 30 mM NaCl pH 8.0. A gradient was used to purify β2GPI between 30 mM NaCl and 350 mM NaCl starting at 0% and finishing at 100% across 1 hour, 5 ml fractions were collected and checked for β2GPI by SDS-PAGE. Samples were then quickly dialysed (centrifugal concentration) or diluted as appropriate to 50 mM sodium acetate, 50 mM NaCl, pH 4.5 (Buffer A), and loaded on a 5 ml SPHP column (Cytiva, Buckingham, UK). Samples were eluted across a gradient of 40-60% Buffer B (50 mM Sodium acetate, 650 mM NaCl, pH 5.3) across 1 hour, peaks were checked for β2GPI by SDS-PAGE. Peaks containing β2GPI were pooled and purified by size exclusion chromatography (16/600, Superdex 200) in phosphate saline buffer (PBS) using a single isocratic wash. Samples were quantified by bicinchoninic acid assay using a bovine serum albumin standard curve.

### Cleavage of β2GPI

Purified β2GPI was cleaved overnight using plasmin (Cambridge Biosciences, Cambridge, UK) at a 1:1 ratio in cleavage buffer (100 mM Tris, 0.02 M NaCl, 0.3 mM CaCl_2_, pH 7.5). Volumes were made up to 500 μl and left rotating overnight at 37 °C. Cleavage was confirmed through reduction studies. 10 μl of reaction mix was incubated with 0.05 M Tris(2-carboxyethyl)phosphine hydrochloride or 0.1 M dithiothreitol for 15 minutes at 95 °C before being run on a 4-12% SDS-PAGE BOLT gel (Invitrogen, Waltham Massachusetts, USA) for 32 minutes at 165 volts. Samples which did not alter their migration under reduction were confirmed to be successfully cleaved.

### Dynamic Light Scattering

The hydrodynamic diameter of β2GPI at 0.1 mg/ml was measured using a Zetasizer Nano-ZS (Malvern Instruments, Herrenberg, Germany). Samples were prepared by filtration (0.2 μM) and centrifugation (16,000 × g, 10 minutes) to reduce potential aggregate formation. Measurements were taken with a detector angle of 90° and a refractive index of 0.01. The pedestal height was allowed to float during measurements ensuring maximal signal. A total of three measurements per protein (six runs with a duration of 10 s per measurement) were obtained and data averaged. Data were analysed using Microsoft Excel and statistical differences were derived using GraphPad Prism 7.0.

### Small Angle X-ray Scattering

X-ray scattering data were obtained during a single beam session on Instrument B21 at the Diamond Light Source at the Harwell Science and Innovation Campus in Oxfordshire, operating with a ring energy of 3 GeV and a beamline operational energy of 12.4 keV (47). A PILATUS 2M detector with a resolution of 1475 × 1679 pixels (pixel size of 172 × 172 μm) was used with a sample-to-detector distance of 4.01 m giving a Q range from 0.04 to 4 nm^−1^ (where Q = 4 π sin θ/λ; 2θ = scattering angle; λ = wavelength). The β2GPI samples, at concentrations between 1.5 mg/ml and 0.1 mg/ml, in buffer were loaded onto a 96-well plate that was placed into an EMBL Arinax sample holder (48, 49). This measurement condition showed the β2GPI molecule as a hydrated structure in a high positive solute– solvent contrast (18). An automatic sampler injected 30 μl of sample from the well plate into a temperature-controlled quartz cell capillary with a diameter of 1.5 mm. Datasets of 30 frames with a frame exposure time of 1 s each were acquired in duplicate as a control of reproducibility. Checks during data acquisition confirmed the absence of radiation damage. Buffer subtraction was carried out automatically. Analysis of Guinier, P(r) and Kratky plots were carried out using ScatterIV and GraphPad Prism 7.0.

### Molecular Dynamics

The crystal structure coordinates of β2GPI (PDB ID: 1c1z) were prepared for molecular dynamics using Glycan Reader and Modeler (29–32) at the CHARMM-GUI website (http://www.charmm-gui.org/). Protonable residues were edited on CHARMM-GUI. Modifications to take account of the plasmin clipping of β2GPI were carried out *in silico* through the deletion of amino acids 318-326 in Pymol. Both plasmin-clipped and healthy control (non-clipped) Β2GPI had full glycan chains added in accordance with the four biantennary glycans detected by Kondo et al (2009).

The TIP3P model was used for explicit water molecules. The cubic system size was determined to give at least 10 Å from the protein in each axis, and 0.15 M NaCl was added. The CHARMM36 force field was used (33,34), and all calculations were performed at 303.15 K. The particle mesh Ewald algorithm was applied to calculate electrostatic forces, and the van der Waals interactions were smoothly switched off at 10 Å by a force-switching function (35). A time step of 2 fs was used in all simulations. Initially, each system was shortly equilibrated in constant particle number, volume, and temperature (NVT) condition using CHARMM36 (36). To assure gradual equilibration of the system, positional restraints for backbone and side chain heavy atoms were applied and the restraint forces were gradually reduced during the equilibration. Each system was further simulated for 100 ns using the CHARMM36 force field on the high-performance cluster, Kathleen, at University College London using NAMD (37). For the production NPT simulation, the Langevin coupling coefficient was set to 1 ps^−1^ and a Nosé-Hoover Langevin-piston (38,39) was used to maintain constant pressure (1 bar) with a piston period of 50 fs and a piston decay of 25 fs. The time step was 2 fs and trajectories were saved every 100 ps. The electrostatic interactions were updated every 20 fs. The short-range non-bonded and electrostatic interactions were calculated with a cut off of 12 Å. SHAKE was used to constrain all bonds involving hydrogen atoms. Convergence of the simulations for all systems was checked through the comparison of average RMSD using VMD (40). Each simulation was repeated three times and the data were averaged for analysis. These were carried out on Kathleen (High Performance Computer at UCL, based on Intel Xeon Gold Processors).

### Inhibition ELISA

Inhibition ELISAs were performed as previously described (23). Briefly, serum samples with historical positivity for anti-β2GPI antibodies were tested in an anti-β2GPI ELISA. Maxisorb plates (ThermoFischer, Waltham Massachusetts, USA) were coated with 2 μg/ml of β2GPI (Enzyme Research Laboratories, Swansea, UK) overnight at 4 °C before being blocked with 2% BSA/PBS (Sigma, St Louis, Massachusetts, USA) for 1 hour at 37 °C (150 μl per well). Plates were washed with PBS Tween (0.1%) three times before application of serum at a dilution of 1:50 in 1% bovine serum albumin in PBS for 1 hour at room temperature (50 μl per well). Plates were again washed and anti-human IgG conjugated to horse radish peroxidase (HRP, Gillingham, Sigma) was applied at a titration of 1:2000. The plate was again incubated at room temperature for 1 hour. Plates were washed and 100 μl of 3,3’, 5,5’-tetramethylbenzidine (KPL, Gaithsburg, Maryland, USA) added and incubated for 15 minutes at room temperature before being stopped with 100 μl of stop solution (1N, KPL, Gaithsburg, Maryland, USA). Plates were read by absorbance at 450 nm, in a plate reader (Tecan Infinite PRO+, Tecan, Männedorf, Switzerland). Inhibition assays were performed identically with the exception of a two-hour pre-incubation of serum samples with either healthy control (non-clipped) β2GPI or plasmin-clipped β2GPI at room temperature before application of sample to the plate. Inhibition was calculated by comparing samples in the absence and in the presence of inhibitor on a single plate.

### Native PAGE

Non-cleaved protein was loaded at 0.6µg, 1.2 µg, 2.4 µg and 5µg in Native loading buffer (Invitrogen). Plasmin-clipped β2GPI was loaded at 5µg diluted in native loading buffer (Invitrogen). Samples were loaded into a 10% Tris-Glycine NATIVE Page Gel (Invitrogen) and run in Tris-Glycine NATIVE Running Buffer at 150v, variable milliamps for 5 hours at room temperature. Gels transferred to PVDF membranes using the BioRad Turboblotter (7 minutes, 10V Mixed Protocol) and blocked with 10% skimmed milk in PBST (0.5%). Membrane was exposed for fluorescence (Thermofisher iBright) for Cy3 and Cy5. Membranes were then incubated with anti-B2GPI antibody (PA-1-74015, Thermofisher) overnight. Membranes were washed with 10% milk in 0.5% PBST for 3 hour (3×20 minutes) and then incubated for 1 hour rolling at room temperature with anti-goat antibody conjugated to HRP (1:1000, DAKO). Finally, membranes were washed with PBST (0.5%) and exposed using Amersham Prime ECL (Amersham) on an Amersham ImageQuant 800 (Amersham, GE Healthcare).

### Fluorescent Labelling and flow cytometry

Glycan labelling of β2GPI was carried out using the N Glycan Labelling Kit (BioTechne). Non-clipped B2GPI was labelled using 1µl Neuramidinase, 1µl StGal6 and 1µl of label per 5µg of protein, incubated at 37°C for 1 hour. Clipped β2GPI was labelled using 2µl Neuramidinase, 2µl StGal6 and 5µl of label per 5µg of protein, incubated at 37°C for 1 hour. Both labelled proteins were stored at +4°C in the dark. Plasmin-clipped β2GPI was labelled with Cy3 whilst non-clipped was labelled with Cy5. Labelled clipped and non-clipped β2GPI were incubated with human umbilical vein endothelial cells (HUVEC) for 2 hours at 37°C. Cells were plated at 500,000/well with 2μg of fluorescently labelled β2GPI. Cells were incubated with Live/Dead Blue viability Dye (ThermoFisher) for 20 minutes in the dark at room temperature. Cells were then stained with CD105-BV421 (BioLegend, 43A3) for 30 minutes at 4oC to allow identification of HUVEC, then incubated in fixation buffer (ThermoFisher) for 15 minutes at 4oC in the dark. Percentages of cells positive for β2GPI were then compared between plasmin-clipped β2GPI and non-clipped β2GPI. Samples were acquired on a Fortessa X20 flow cytometer (BD). Data were analysed by FlowJo (v10).

### Ion Mobility Mass Spectrometry

Ion Mobility Mass Spectrometry was carried out in the Thalassinos Laboratory at UCL. Protein samples were buffer exchanged into 10 mM ammonium acetate using an Amicon 30 kDa MWCO filter for analysis on a SELECT SERIES Cyclic IMS QToF (Waters Corp.) (41,42). Samples were direct infused into the instrument at a concentration of 2.5 μM using nano electrospray capillaries prepared in house using a Flaming-Brown P97 micropipette puller, and gold-coated with a Quorum Q150RS sputter coater. Data were processed using UniDec and MassLynx v4.2.

## RESULTS

### Validation of cleavage of β2GPI by plasmin

β2GPI was cleaved successfully overnight (16 hours, 1:1 ratio) yielding approx. 50% cleaved protein. Confirmation of cleavage was achieved by SDS-PAGE (Figure 1A) and native PAGE/Western Blot (Figure 1B). In the native gel/western the plasmin clipped protein forms a wider smear with a higher mobility under native conditions, suggesting that it is more flexible and potentially more compact compared to the non-cleaved linear structure. The affected sites from cleavage are modelled in Figure 2. Protein was purified from plasmin mix using ion exchange chromatography, showing a shift in affinity under cation exchange conditions, eluting at a lower conductivity. This confirmed cleavage and enabled good separation of cleaved and non-cleaved β2GPI (not shown).

**Figure 1:**
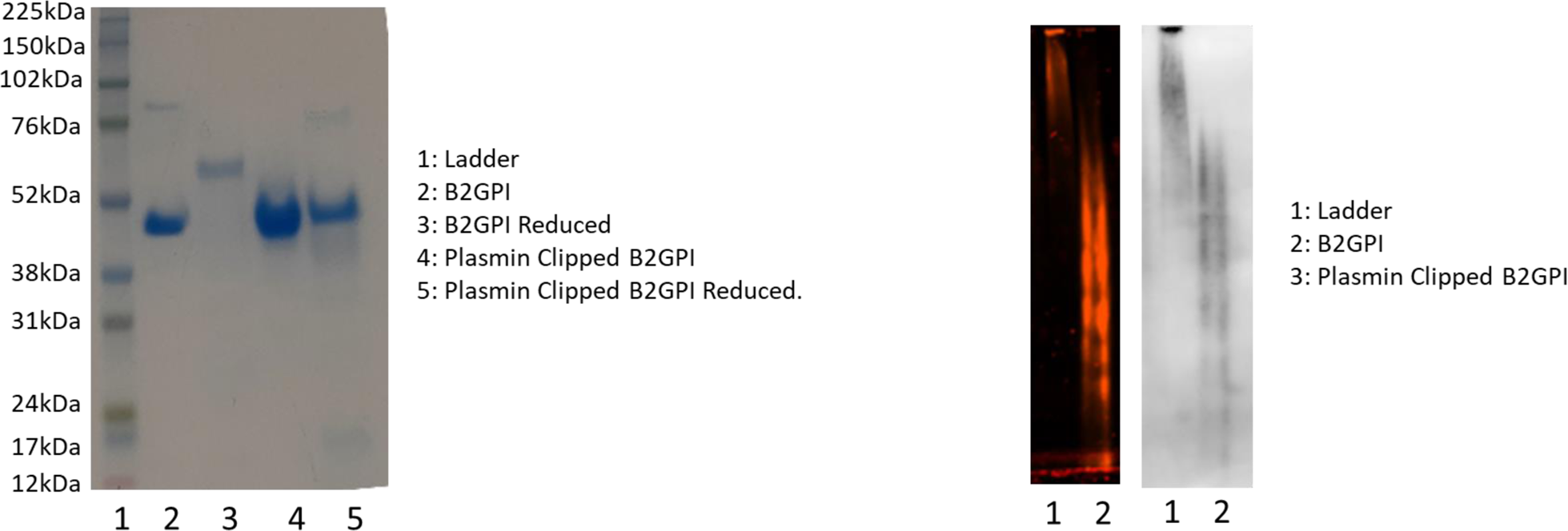
Production of plasmin clipped B2GPI. Batches of protein were cleaved and confirmed for cleavage by reduction SDS PAGE analysis. Whole B2GPI when reduced has reduced mobility on an SDS PAGE Gel (lane 3) however, after cleavage this reduced mobility is no longer seen (lane 5). Panel B shows the fluorescently labelled B2GPI and Plasmin clipped B2GPI by Native Gel with the lower smear of structures representing the plasmin clipped B2GPI and the higher smear representing non-clipped B2GPI. This was then probed with an ant-B2GPI antibody, showing specificity on the right, highlighting a different structure in Plasmin Clipped B2GPI

**Figure 2:**
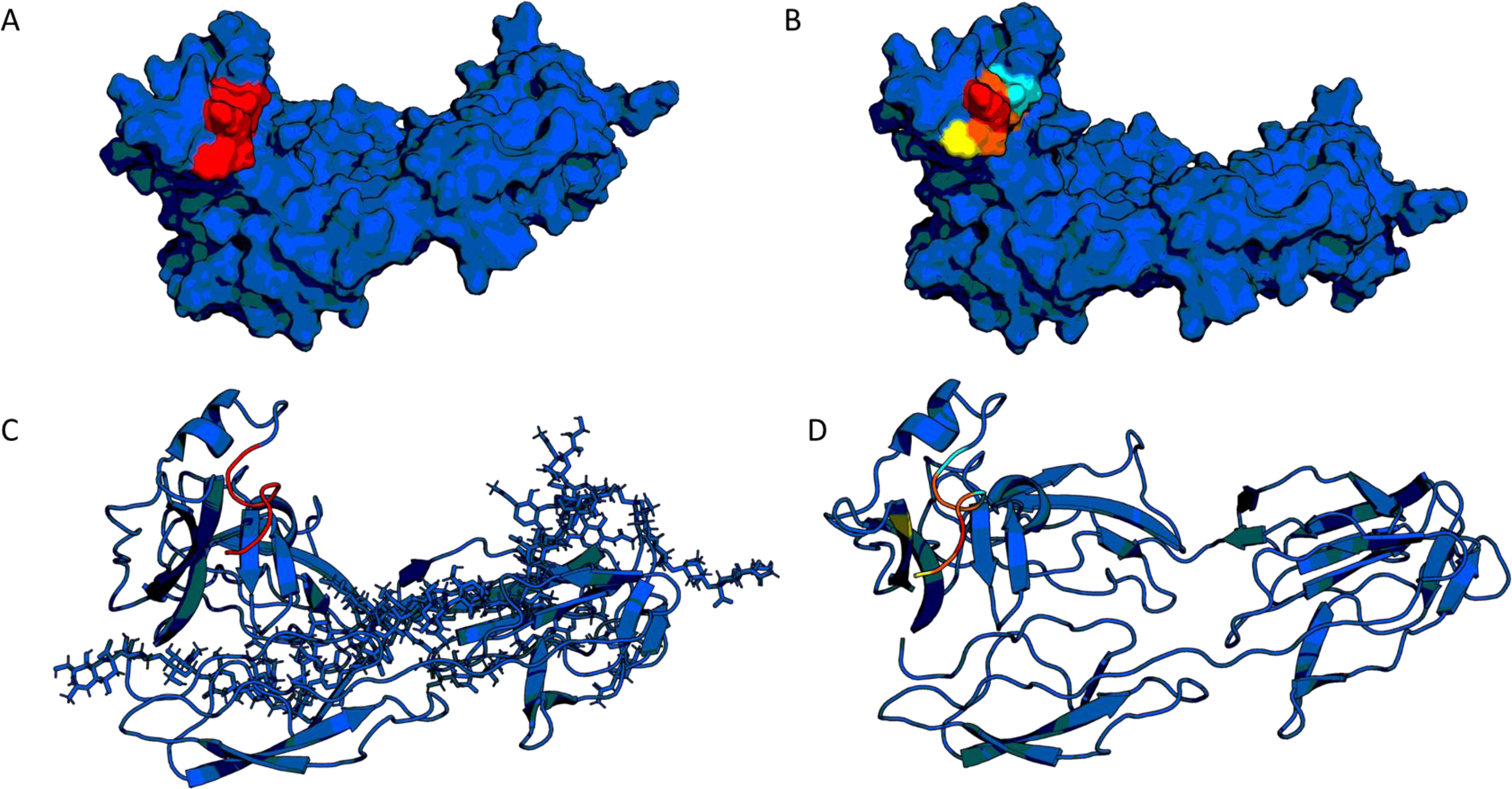
Altered Charge and Interactions. Plasmin cleavage removes the terminal 8 amino acids, leaving a truncated protein, the cleaved peptide being highlighted in red in panel A and C (surface model and ribbon model respectively). This peptide consists of 3 highly charged amino acids highlighted in panel B: one Lysine (Red) two Aspartic acids (Cyan), whilst also containing a cysteine which forms part of an allosteric disulphide with the neighbouring beta-sheet (yellow). The partner cysteine is internal to the structure, as such it has been highlighted in the ribbon model in yellow. Allosteric disulphides are associated with significant structural shift, as such the loss of the disulphide coupled with the alteration in local charge suggest it is likely B2GPI undergoes a significant structural change post plasmin cleavage.

### Native IMMS Data Confirm Altered Structure

Analysis of the non-clipped and clipped β2GPI by means of native ion mobility mass spectrometry revealed differences between the two samples, both in the mass spectrometry and ion mobility dimensions. As seen in Figure 3A, multiple peaks can be seen for each charge state due to glycosylation. While this is true for both samples, for the plasmin clipped sample there is a shift toward the lower charge states being more prominent which is indicative of a more compact / folded conformation. This is shown further in the arrival time distributions obtained from the IM analysis. IM separates ions based on their interaction with a buffer gas as they travel through a mobility cell under the influence of a weak electric field. The time an ion takes to traverse the IM cell is related to its charge and rotationally averaged collision cross section (CCS or Ω), the latter being a physical quantity related to the overall shape (43). For ions of the same charge state, the more compact species will therefore have a faster arrival time distribution (ATD) than an extended one. While the plasmin-clipped protein has one peak in the ATD, the non-clipped version has an additional, later arriving peak illustrating that the protein co-exists in two broad conformers, a compact and an extended one.

**Figure 3:**
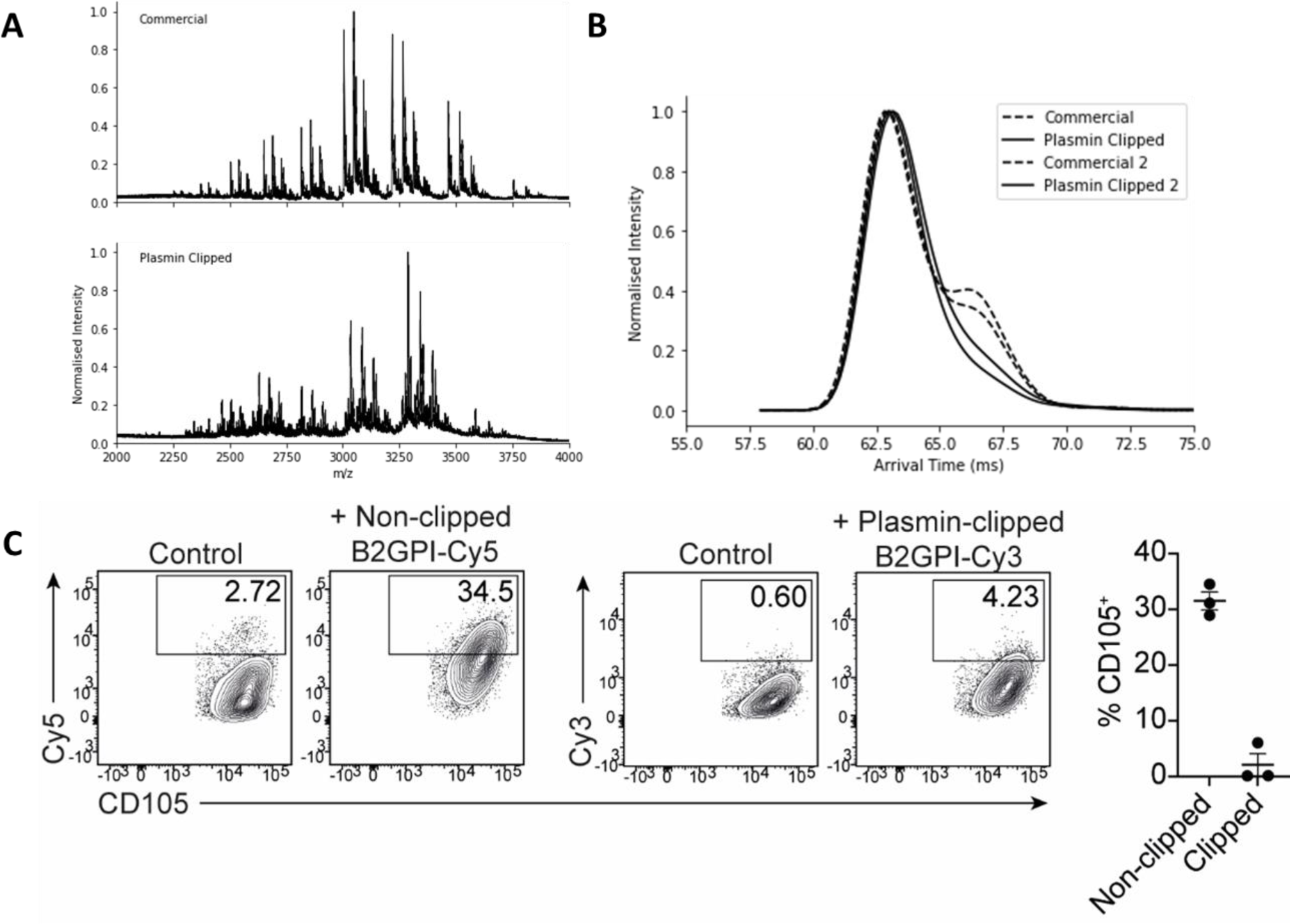
Ion Mobility Mass Spectrometry and FACs analysis. Panel A shows the spectra of both the non-clipped B2GPI (top) and the plasmin clipped B2GPI (bottom), as can be seen both retain a number of glycoisoforms, however, the distribution of charge states is different in the Plasmin Clipped B2GPI suggesting a difference and a more compact conformation. Further to this in B, Ion Mobility data shows in non-clipped B2GPI that there two conformational families, with the dominant conformer corresponding to the earlier peak. In contrast plasmin clipped B2GPI shows a single dominant conformational family, suggesting that cleavage results in the loss of the extended form. Plasmin Clipped B2GPI is known to lose its ability to bind to cell surfaces, as such we conducted FACS with fluorescently labelled B2GPI (clipped and non-clipped) on HUVEC cells. As can be seen, non-clipped B2GPI (Cy5 labelled) bound to approximately 3-35% of cells (2ug B2GPI, 500,000 cells) whilst plasmin clipped B2GPI bound between 0 and 4.5% of cells, significantly less.

### Flow Cytometry Shows Consistent Activity

Non-cleaved and plasmin-clipped β2GPI were conjugated to Cy5 and Cy3 respectively and incubated with human umbilical vein endothelial cells (HUVEC) for 2 hours at 37°C. HUVEC were stained with CD105-BV421 for identification by flow cytometry. Significantly higher frequencies of CD105^+^ HUVEC (P<0.05) were positive for non-clipped β2GPI (30%) compared to plasmin-clipped β2GPI (<5%) consistent with findings from other studies. This confirmed that cleavage has been successful and suggested that fluorophore labelling has not altered the physical function of the protein facilitating binding of the protein to cellular surfaces.

### ELISA assays

Activities of non-cleaved and plasmin-clipped β2GPI were measured using in-solution inhibition assays. In Figure 4A the y-axis shows inhibition, defined as the percentage reduction in binding in the presence of inhibitor (clipped or non-clipped β2GPI) compared to absence of inhibitor. The mean inhibition was 14.9% (SD 8.9%) for plasmin-clipped β2GPI compared to 9.4% (SD 9.1%) for non-clipped β2GPI (p=0.056). This suggests that the structural alteration may influence the ability of β2GPI to bind pathogenic antibodies in favour of higher binding for plasmin-clipped β2GPI, potentially due to changes in surface exposure.

**Figure 4:**
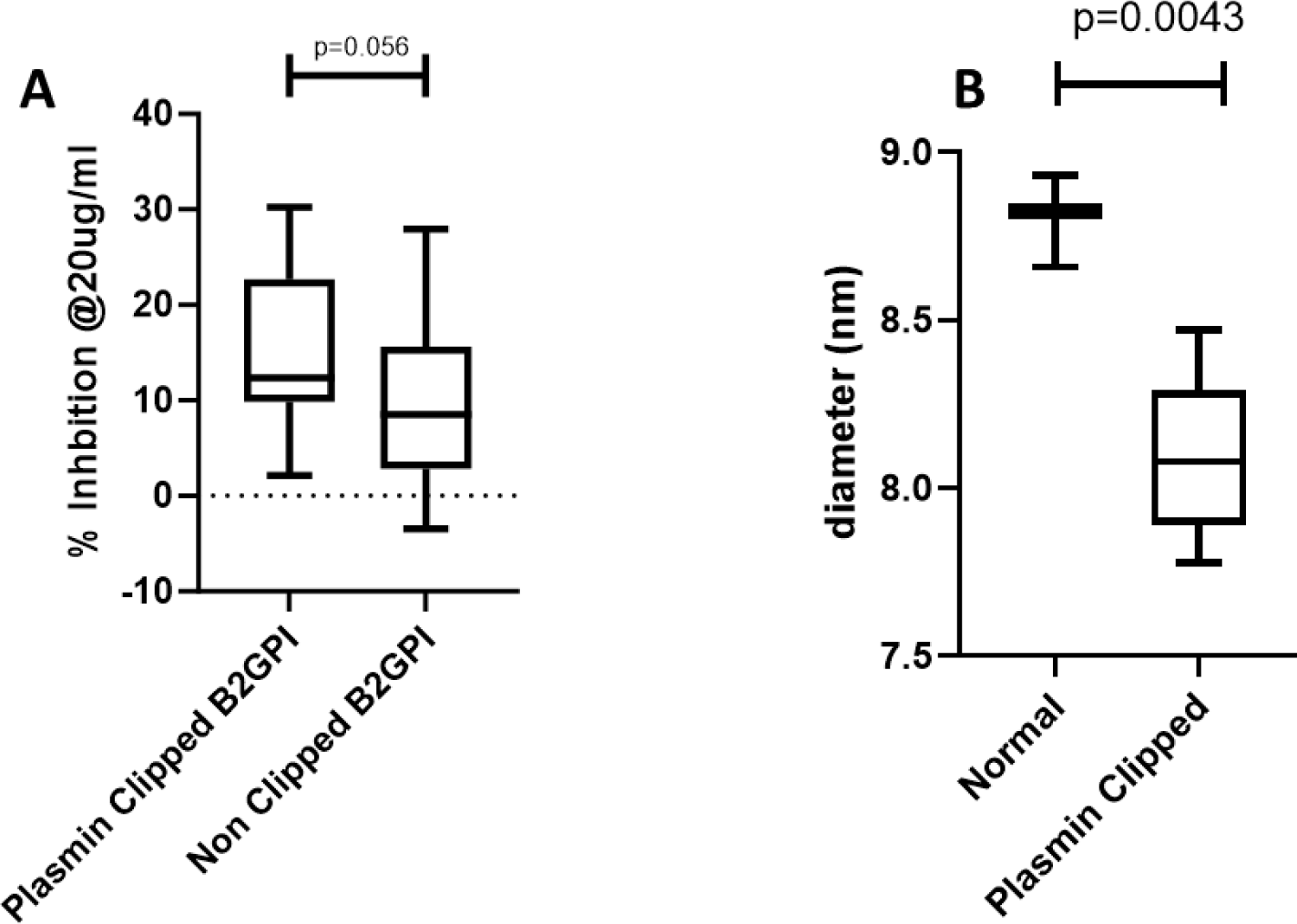
Activity and Structure. To test the activity of the plasmin clipped B2GPI vs the non clipped B2GPI we carried out competitive inhibitions using patient serum. Anti-B2GPI positive serum was pre-incubated with 20ug/ml of either Plasmin Clipped B2GPI or non-clipped B2GPI before exposure to a pre-coated B2GPI ELISA plate, a loss of binding to the plate was defined as an increase in inhibition by the in solution inhibitors. As can be seen in A, plasmin clipped B2GPI showed greater inhibition than non – clipped suggesting a different structure with a greater exposure of epitopes. Panel B shows the dynamic light scattering profile of both the plasmin clipped and non-clipped B2GPI, as can be seen the non-clipped B2GPI has a higher dynamic radius, suggesting a larger, defined structure whilst the clipped B2GPI shows a larger variation and a smaller size suggesting the structure of plasmin clipped B2GPI is more flexible, and smaller than non-clipped B2GPI.

### Altered structural profiles after plasmin cleavage revealed by DLS

DLS measurements showed a significant difference (p=0.039) in hydrodynamic radius between the plasmin-clipped β2GPI (n=6, 7.8 nm) and non-clipped β2GPI (n=3, 8.8 nm), Figure 4B. This difference of approximately 1nm may be explained by a slightly different structural profile, with a smaller hydrodynamic radius accounting for a more compact form. This corresponds to the changes seen by Native PAGE.

### SAXS comparison of plasmin-clipped and healthy control (HC, non-clipped) β2GPI

Plasmin-clipped and non clipped β2GPI were imaged at the Diamond Light Source Facility in the Harwell science campus (UK) for SAXS analysis. A range of concentrations from 1.5 mg/ml to 0.2 mg/ml were imaged with optimal comparative data being obtained for concentrations of 1 mg/ml. Buffer was subtracted automatically post-normalisation, and no difference in intensity was seen between the two proteins (Figure 5A).

**Figure 5:**
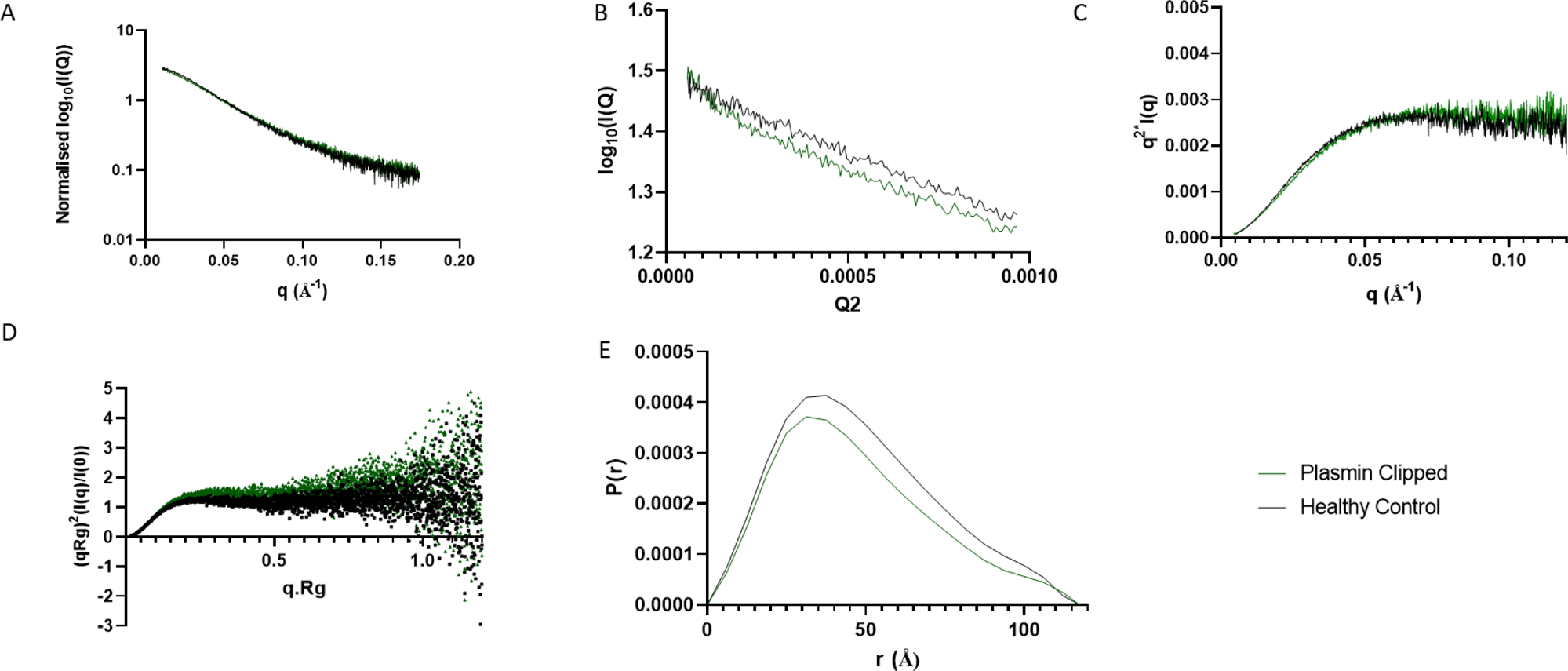
Small Angle X-ray Scattering of Plasmin Clipped (Green) and Healthy Control (non-clipped) (Black) purified B2GPI. Plot A shows a Kratky plot of both proteins, the peaks are at a similar point suggesting they are both folded similarly, however, the green line maintaining a higher plateau may suggest increased flexibility in the Plasmin Clipped B2GPI. A normalised intensity plot (B) shows no significant difference between the proteins whilst the Guirnier fitting (C) shows the P(r) plot, with a characteristic high peak with extended tail shown in more rod shape structures, whilst the uneven shoulder shows the multi-domain nature of the protein. Panel D shows the Guirnier fit whilst E shows a dimentionless Kratky plot, again the plasmin clipped has a more defined peak and trends above the healthy control (non-clipped) demonstrating the increased flexibility the cleavage facilitates.

Guinier analysis was carried out on both HC and plasmin-clipped β2GPI curves identifying a single linear region (Figure 5B). The Q. R_G_ limits were similar between both proteins with the HC Q-range from 0.446 Å^−2^ to 1.287 Å^−2^ whilst plasmin-clipped was 0.571 Å^−2^ to 1.297 Å^−2^. These ranges were selected as they gave the most linear fit and the inclusion of values up to a Q. R_G_ of 1.3 is in line to include globular, disk- and rod-shaped objects. The Rg was calculated to have values of 40.9 Å and 40.3 Å, respectively for HC and plasmin-clipped β2GPI.

Pair-distance distribution function analysis P(r) was used to provide structural information in real space, where the largest Q value was limited to 0.13 nm^−1^. D_max_ was set to 118 nm for both proteins by trial and error, giving the smoothest descent to zero and some slight aggregation was seen in the plasmin-clipped sample at very low Q values, however, this region was before the Q values used in the Guinier analysis. R_G_ values from the P(r) analysis were smaller than for the Guinier analysis with healthy control (non-clipped) calculated at 38.4 Å whilst plasmin-clipped was calculated at 37.6 Å. These values are within 10% of the Guinier analysis values and predicts a smaller R_G_ for plasmin-clipped β2GPI, which is consistent with the DLS data. The peaks seen in the P(r) analysis are formed by the most frequently occurring atomic distances within the structure, the dominant peak in both healthy control (non-clipped) and plasmin-clipped β2GPI occurs between 30 and 40 Å, whilst the shape of the peak suggests a long rod shape. The maximum length of the protein (L) was calculated using the P(r) function identifying the shape as a long rod and solving the equation for L (44):

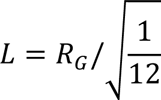

This gave a length of 132.8 Å for HC β2GPI and 130.2 Å for plasmin-clipped β2GPI, again suggesting a smaller size for plasmin-clipped β2GPI. Analysis of Kratky plots (Figure 5C) showed a more defined maximum for the plasmin-clipped protein (green) compared to healthy control (non-clipped) β2GPI (black), whilst a dimensionless Kratky plot showed both proteins had a multidomain profile (Figure 5D). This suggested a less linear, although not a fully compact globular protein form. This may also point to increased flexibility in the plasmin-clipped β2GPI.

Further to this, the P(r) function (Figure 5E) showed the characteristics of an elongated protein, with an initially well-defined peak before a tail around 75 Å. Although both samples showed this profile, a difference was seen in the height of the peaks, suggesting that although the two proteins were extended, they were non-identical in structure.

### Molecular Dynamics

To generate potential model conformations that could explain the SAXS data, three 100 ns repeats of molecular dynamics simulations were run. These were based on the crystal structure of β2GPI (PDB ID: 1c1z) and used a simulated temperature of 313 K. These simulations were then used as sources of β2GPI structures to generate theoretical SAXS curves, to identify those conformers which fit best to the SAXS data (45,46). SASSIE was used to generate the theoretical curves from structures, which were then analysed for best fit to experimental data using a Chi Squared filter. The top 100 structures were identified and plotted in Figure 6. All structures associated with healthy control (non-clipped) β2GPI (bottom) had a smaller range of radius of gyration (R_G_) than for the plasmin-clipped (top). The top 100 structures, highlighted in red, were significantly different in their R_G_ values (p=0.0001) with those for healthy control (non-clipped) ranging from 42.3 to 46.7 nm, whilst those for plasmin-clipped ranged from 38.9 to 43.6 nm. This reflects the smaller values for plasmin-clipped seen in the DLS data.

**Figure 6:**
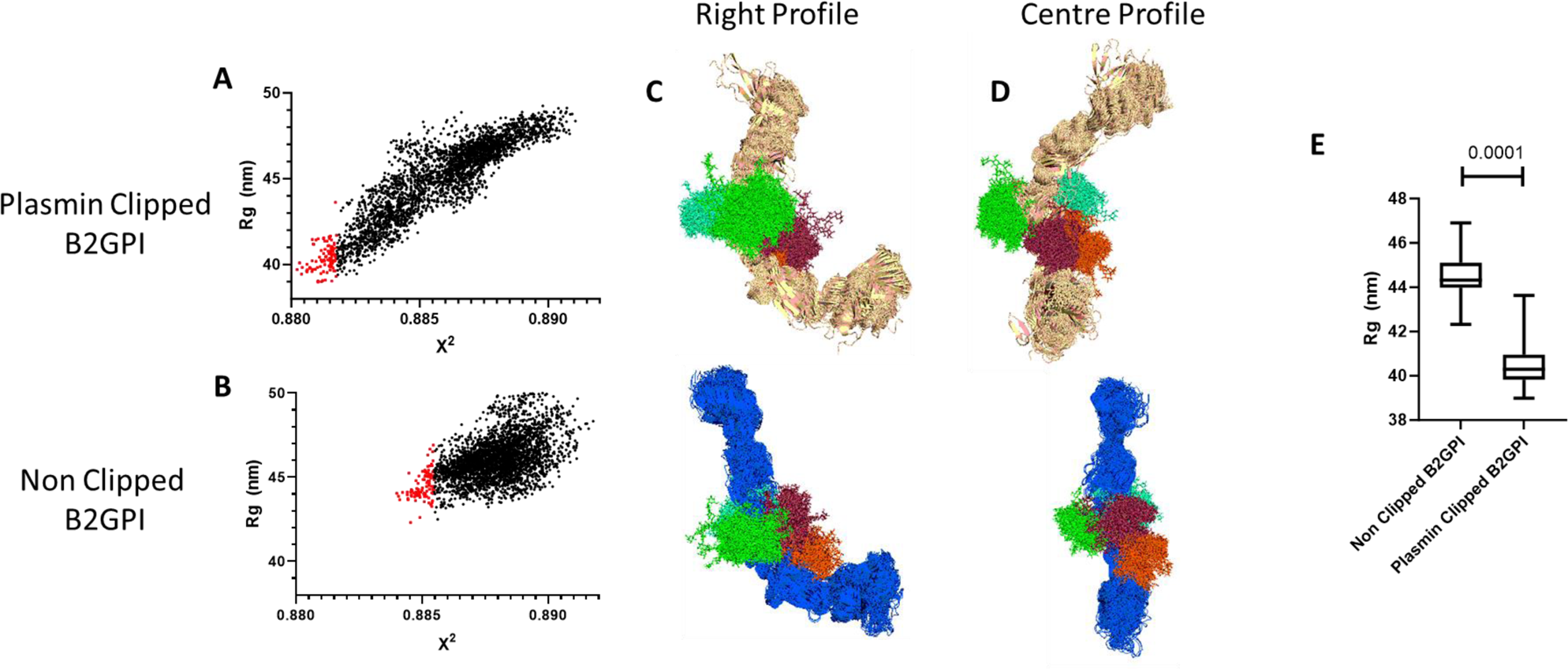
Analysing the output from the SAXS data using Molecular Dynamics using >3000 individual frames, we identified the top 100 fits for each structure from their respective simulation. Plotting the Rg vs X2 highlighted that the non clipped B2GPI had structures with a higher Rg (**A,B**). Further to this, visualising the structures in Pymol highlighted the plasmin clipped B2GPI has a significantly different structure, with the 1^st^ 2 domains (DI, DII) rotated **(C,D)**. Plotting the best fitting 100 frames for both non clipped and clipped B2GPI showed a significantly higher Rg for non-clipped compared to plasmin clipped **(E**) confirming what was seen by SDS PAGE and IMMS.

These top 100 hits were aligned and overlaid to evaluate whether there was a single consistent structure associated with either the healthy control (non-clipped) or plasmin-clipped β2GPI. As can be seen in Figure 6, healthy control (non-clipped) (blue) had a consistently extended J-shaped linear form of the structure, as has been observed previously by a number of methods including crystal structures (5), AFM (47) and SAXS (48). By contrast, the plasmin-clipped β2GPI (Beige) consistently gave a novel S-shaped linear form of the structure, which was therefore more compact than the J-form. When overlaid with each other and aligned based on the 3^rd^ and 4^th^ domains (DIII, DIV), there was a clear and significant divergence in the position of the 1^st^ 2 domains (DI, DII) (Figure 6C/D). The single best-fit models were then overlaid to show more clearly the difference (Figure 6C/D) between the two structures. The R_G_ was also significantly different with smaller values seen in the plasmin-clipped, as shown in DLS confirming the SAXS and MD were consistent with other laboratory processes (Figure 6E).

Applying Principal Component Analysis (PCA) to the top 100 conformations of each of healthy control (non-clipped) and plasmin-clipped β2GPI showed a clear difference, with their structures clustering independently when using the hclust function in R (hierarchical, complete linkage) (Figure 7C) this was further demonstrated using aligned structures of best fit in Figure 7A and Figure 7B. To avoid clustering being influenced by the removal of 8 amino acids, all frames were trimmed to contain identical constructs, the proteins aligned using all carbon atoms before the PCA was carried out. Further to this, highlighting the DI epitope on a space model (Figure 7E) shows that the epitope has a potentially very different solvent exposure in the plasmin-clipped structure. Analysis of the entire simulations independently of the SAXS data showed a significantly increased RMSD for the plasmin-clipped β2GPI over time. At the amino acid level, the regions of greatest movement as measured by RMSD, were found in the 1^st^ and 5^th^ domains (DI, DV, Figure 8). This suggests that the switch into this novel S-shaped structure after cleavage by plasmin, may be associated with an overall increase in flexibility within the protein as the terminal domains are both moving in opposition to one another. Furthermore, when clustering all conformations and not just the frames extracted from SAXS analysis, and investigating the effect of each amino acid on the principal components (PCs), the 5^th^ domain (DV) drives the separation in the 1^st^ 5 PCs (Supplementary Figure 1) suggesting this is driving the most significant structural change.

**Figure 7:**
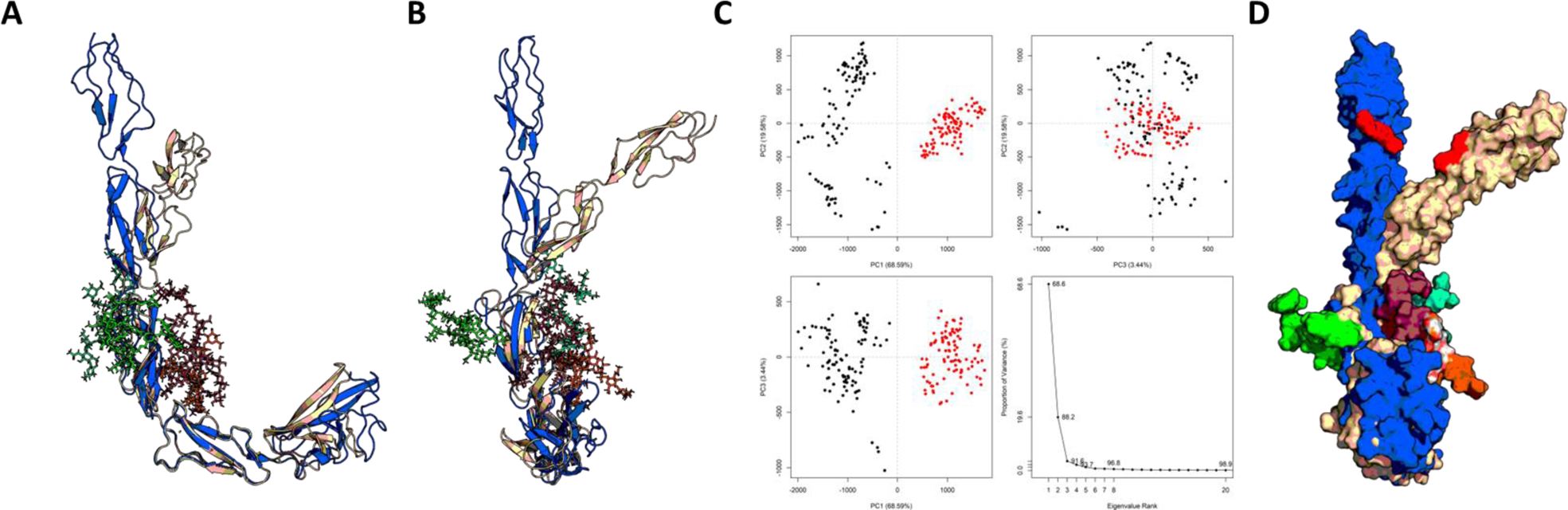
Alignment of the best fitting structures from the molecular dynamics simulations by domain IV (which shows the least difference) highlights the alterations in the orientation of DI and DII **(A & B).** When combining the top 100 frames of each of the clipped and non clipped B2GPI and conducting a PCA analysis, it shows that the two sets of structures are significantly different with PC1 accounting for almost 70% of differences whilst PC2 accounts for another 19% **(C)** with the black dots being plasmin clipped and the red dots non-clipped B2GPI, also showcasing the high variability in structures in the plasmin clipped compared to the non-clipped B2GPI. Finally two space models of the best fits are overlaid with the R39-G43 epitope highlights, as can be seen it not simply a case of the protein bending but in the plasmin clipped B2GPI it also rotates exposing the epitope differently, perhaps explaining the increased binding of antibodies in serum to the plasmin clipped B2GPI.

**Figure 8:**
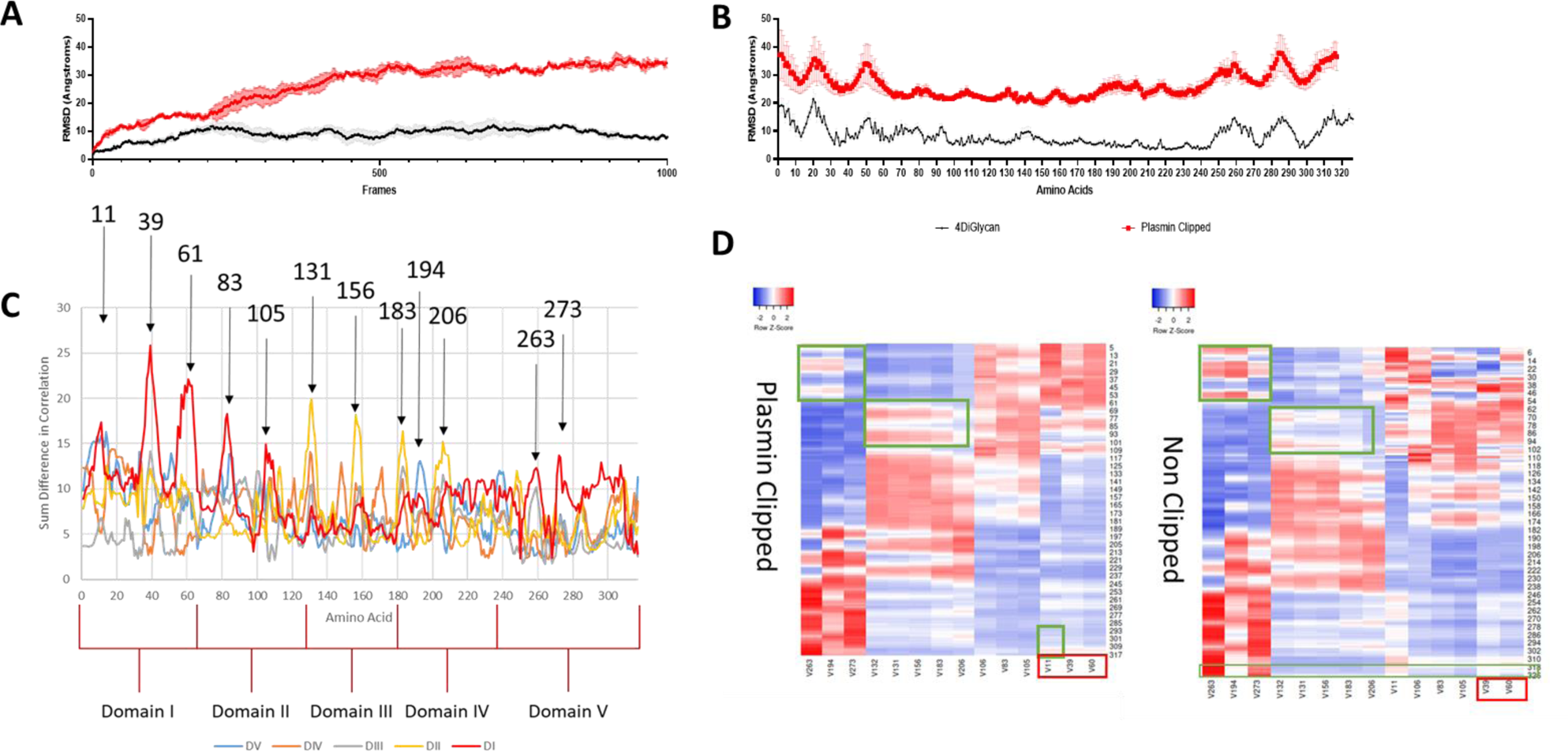
Molecular simulation of B2GPI. Panel A and B show the RMSD of non-clipped B2GPI across the simulation compared to plasmin clipped B2GPI **(A)** and the same comparison by amino acid **(B).** As can be seen in both measures plasmin clipped B2GPI has a far greater flexibility and movement suggesting a significant structural change occurs during cleavage. This corroborates the wet lab work suggesting a different structure and higher variation in structure. Further to this an analysis of intra-protein networking showed that significant differences were seen across multiple domains in the DCCM matrixes, this is demonstrated in **(C)** where sum differences in correlation per amino acid vs domains are plotted. As can be seen the red line (DI) shows the most significant changes in movement correlations with both itself and long range to DV, similar the yellow line (DII) shows significant alterations in correlation with the 3^rd^ (DIII) and 4^th^ (DIV) domain. A number of amino acids are picked out as high points for variation, pointing to a pathway throughout the protein effected by the modification. Plotting these variations on a heatmap for their correlation with other domains and themselves **(D)** reveals a backbone of amino acids which correlate with domains throughout the protein. Interestingly, significant alterations are seen in how these amino acids signal together, the green boxes show areas where correlations are altered, with more correlation between V132-V206 with DII in the Plasmin Clipped, whereas these are less strongly corelated in the non-clipped model. Similarly, DI correlates with V19, 273 and 194 strongly in the non-clipped model but this is largely lost in the non-clipped model suggesting this long distance relationship is altered by clipping. The grouping of the amino acids themselves is changed too, with V11 clustering more with DI in the plasmin clipped model compared to the non-clipped which shows more correlation to later amino acids (V106-206). This lends credence to the idea that the cleavage of the terminal amino acids in Plasmin Clipped leads to an alteration in the movement of the 1^st^ and 2^nd^ domains (DI, DII), as seen in the previous experiments.

Plotting the other variables (RMSD/ R_G_) for the top 100 fitted structures from the SAXS analysis identifies structures with a high RMSD and a low R_G_ suggesting they are flexible but compact, which links with the initial data from the DLS and Native PAGE (Supplementary Figure 1C). Furthermore these structures are found in the 1^st^ cluster predominantly which occurs early in the simulations, suggesting that they are quickly adopted and potentially therefore low energy.

## DISCUSSION

The role of plasmin-clipped β2GPI is still a mystery in the context of APS with little known about its structure and its function. It has been shown that this β2GPI cleavage can alter binding to negative surfaces and alter the role β2GPI plays in plasminogen generation (12,49). However, its role in autoantibody binding has not been assessed.

The structure of β2GPI is somewhat controversial, Agar proposed a linear (J) and a circular (O) form based on electron microscopy methods (1). More recently, other groups (3) have postulated that an S form exists, though it has not been proven. Most recently the group of Pozzi have imaged β2GPI after molecular mutation to remove a disulphide bond in the 5^th^ domain (DV) showing a difference in structure (50). Finally, Buchholz et al. have shown structural change when lysines undergo post translational modification *in vitro* resulting in an altering of the linear-circular equilibrium (9). However, all of these modifications are *in vitro* and may not reflect exactly what occurs in patients. Plasmin cleavage has been confirmed to occur *in vivo* and is theorised to occur more frequently within APS patients. Given the role of clotting in disease progression and the role of β2GPI in thrombosis and complement activation, which has been shown to be required for clotting in APS patients (2) it is fair to suggest plasmin-clipped β2GPI does exist in APS patients *in vivo* and the lack of proof is a sign of the difficulties in assaying the molecule.

This study is the first to apply biophysical and computational methods to plasmin-clipped β2GPI and has revealed a novel, potentially pathogenic structure of β2GPI. Traditional structural biology methods such as dynamic light scattering suggested a compact variant of the linear structure of β2GPI after plasmin cleavage. This was then corroborated by the SAXS analysis which highlighted structures with smaller radius of gyration, and then revealed the S-shaped structure of plasmin-clipped β2GPI through fitting of the data to MD simulation structures. This structure was associated with an increased ability to bind pathogenic autoantibodies from patients in serum as determined by ELISA, which has never been described previously for plasmin-clipped β2GPI.

The identified novel conformation may also explain the unusual functions of plasmin-clipped β2GPI. As discussed, plasmin-clipped β2GPI can act as an inhibitor of angiogenesis. However, this appears to be an enhancement of an inhibition activity already seen in the non-clipped β2GPI. This enhancement may be due to the change in structural distribution between linear and circular β2GPI, caused by plasmin cleavage. Reed’s work on the potential for increased neonatal cardiac manifestations showed that β2GPI was able to bind and sequester Ro60 when in its non-cleaved for. However, the cleaved peptide generated in plasmin-clipped β2GPI (consisting of 8 amino acids) may contain the sequence required for binding (17). This directly points at conformational change altering the function of the protein. The effect of plasmin cleavage in cerebral infarct in APS patients is important as we have shown increased binding of pathogenic antibodies to plasmin-clipped β2GPI. The mechanism behind this activity is incompletely understood with plasmin-clipped β2GPI binding to Glu-plasminogen, whereas the non-clipped β2GPI cannot. This leads to decreased fibrinolysis and thus, increased clotting, and is hypothesised to be through binding via the neutralised Lysine cluster generated by a structural change first identified by Matsuura et al (51). This structural change in domain V is confirmed in our work with the SAXS model showing significant change in domain V, including around the Lysine rich region.

To conclude, plasmin cleavage of β2GPI resulted in a significant change in structure yielding a novel S-shaped structure capable of increased antibody binding. This structure has increased flexibility and fits well with the literature showing an altered activity in plasmin-clipped Β2GPI compared to healthy control (non-clipped) β2GPI. The demonstration of this new structure is an important step in understanding the functions of β2GPI within the body.

## Data Availability Statement

All data are contained within this manuscript and data are available on request.

## Ethics

Research samples were collected by written informed consent (National Research Ethics Committee-London Hampstead, reference number 12/LO/0373).

## Acknowledgements

We thank Dr Felix Nagel for his insight in discussing the structure of β2GPI during this project, and Dr Nathan Cowieson and Nikul Khunti for excellent user support on Beamline B21 at Diamond. We also thank Stephen McElvaney for his crucial role in logistical facilitation of this study. We also thank Dr Valentina Spiteri for her invaluable insight during drafting.

## Author contributions

T.C.R.M. conceived and coordinated the study, carried out the experiments, and wrote the paper. H.B. recruited the patients and purified the cells and serum from them and conducted Flow Cytometry experiments and analysis. C.L. helped perform the MD simulations and SAXS analyses. X. G. assisted with the SAXS data collection. C.P. developed the original β2GPI inhibition assays and aided in their analysis. A.R. and I.G. provided the ethics permissions and patient samples. H.B. acquired IMMS data under the supervision of K.T. S.J.P. oversaw the SAXS analysis and helped write the manuscript. K.T., M.D. and P.A.D. helped with the study design, analytical development and manuscript drafting.

## Funding and additional information

We thank the Medical Research Foundation for a Fellowship to T. C. R. M. (MRF-057-0004-RG-MCDO-C0800) and a Versus Arthritis Senior Fellowship (ShS/SRF/22977). H.F.B. was supported by an UCB BIOPHARMA SPRL/BBSRC PhD Studentship (BB/P504725/1). C.J.L. was supported by the EPSRC Centre for Doctoral Training in Emergent Macromolecular Therapies (000033549) and IPSEN Bioinnovation (EP/L015218/1). S.J.P. was supported by the CCP-SAS project, a joint EPSRC (EP/K039121/1) and NSF (CHE-1265821) grant. P.D is supported by an EPSRC Grant (EP/P006485/1). Wellcome Collaborative Award in Science (209250/Z/17/Z) to K.T.

## Abbreviations

The abbreviations used are

β2GPI: β2 glycoprotein I
HC: healthy control
MD: molecular dynamics
*R_G_*: radius of gyration
SAXS: small angle X-ray scattering
IEX: Ion Exchange Chromatography
ELISA: Enzyme Linked Immunosorbent Assay

